# Application of an Opioid Use Disorder Cascade of Care in a Large Public Health System

**DOI:** 10.1101/2023.10.19.23297271

**Authors:** Emily Carter, Daniel Schatz, Noah Isaacs, Juan Garcia, Brandy Henry, Noa Krawczyk, Arthur Robin Williams

## Abstract

**Background:** Over the past decade, hospitals and health systems have increasingly adopted interventions to address the needs of patients with substance use disorders. The Opioid Use Disorder (OUD) Cascade of Care provides a framework for organizing and tracking patient health milestones over time, and can assist health systems in identifying areas of intervention to prevent overdose and maximize the impact of evidence-based services for patients with OUD. However, detailed protocols are needed to guide health systems in how to operationalize the OUD Cascade and track outcomes using their systems’ electronic medical records (EMR).

**Objective:** In this paper, we describe the process of operationalizing and implementing the OUD Cascade in one large, urban, public hospital system.

**Methods:** Through this case example, we describe the technical processes around data mining, as well as the decision-making processes, challenges encountered, and lessons learned from compiling patient data and defining stages and outcome measures for the OUD Cascade of Care. The current established framework and process will set the stage for subsequent research studies that quantify and evaluate patient progression through each stage of OUD treatment across the health system and identify target areas for quality improvement initiatives to better engage patients in care and improve health outcomes.

**Results:** The current paper can therefore serve as a primer for other health systems seeking to implement a data-informed approach to guide more efficient care and improved substance use-related outcomes.

**Conclusion:** An OUD Cascade of Care must be tailored to local systems based on inherent data limitations and services design.

## INTRODUCTION

A Cascade of Care is a public health framework for organizing, monitoring, and improving effectiveness of evidence-based care and achieving health milestones. First promoted for HIV/AIDS viral suppression (1), the framework supports tracking health goals across sequential care stages over time. Patient progression within a full cascade of evidence-based services is measured at either individual and population levels to guide intervention development/service delivery, identify disparities, and inform quality improvement. Analogous to evidence-based treatment for HIV infection, medication for opioid use disorder (OUD) (methadone, buprenorphine, and extended-release naltrexone), offers great health benefits, including overall improved quality of life and reductions in: opioid use, overdose, all-cause mortality, and infectious disease transmission (2, 3).

The OUD Cascade of Care assists health systems and other stakeholders in identifying points to maximize impact of evidence-based services on OUD prevalence and overdose. Health systems are using the OUD Cascade of Care to measure system-level OUD treatment engagement and medication for Opioid Use Disorder (MOUD) initiation and retention (4, 5, 6). However, non-standard definitions of stages, challenges in electronic health record (EHR) extraction, and data manipulation/linkage complexities curtail efforts to construct rigorous evaluations of patient progression through the OUD Cascade of Care (7). Successful examples and strategies of how health systems operationalize the OUD Cascade of Care are needed.

The OUD Cascade of Care framework first published by Williams et al. in 2017 (8) and expanded on in 2018 (9) and 2019 (10) defined five sequential stages of treatment for people with OUD: 1) OUD identification (diagnosis); 2) treatment engagement; 3) MOUD initiation; 4) MOUD retention (e.g. for a minimum of 6 months); and 5) Remission. In 2022, Williams et al. (7) published a roadmap for healthcare systems and state agencies developing their own OUD Cascade models, which elaborated on additional design domain considerations beyond stage definitions. Adapted from the HIV work of Haber et al. (11), Williams et al. outlined two domains for Cascade construction: *measurement design* and *scope of staging*.

*Measurement design* refers to four aspects of a cascade of care: 1. Window of observation (longitudinal v. cross-sectional); 2. Single or multiple populations; 3. Denominator-numerator linkage; and 4. Denominator-denominator linkage (7). Cross-sectional analyses allow for the identification of patients that meet the criteria for a specific stage within a specific period of time, while longitudinal analyses follow the same individuals over time as they progress through Cascade stages. A Cascade that follows a single population includes patients from the same defined population, while a multiple population cascade may include patients from different sources reflective of different time periods, clinical settings, or locations. Denominator-numerator linkages use the same population within but not across stages, while denominator-denominator linkages use the same population for all stages (7).

*Scope of staging* refers to breadth and depth, where breadth is the range between the first and last stage, and depth is the number of stages between the first and last stages (7). A broader Cascade could span from the time someone becomes at risk of developing OUD to the time that someone is in remission. For example, patients who take prescription opioids for pain management or use opioids but do not meet the DSM-5 criteria for OUD may fall into an “at risk” stage and may benefit from intervention (12, 10). At the other end of the Cascade, patients who achieve complete abstinence and/or no longer meet the DSM-5 criteria for OUD for 90 days may fall into a “remission” stage (12, 13, 10). A deeper Cascade framework might disaggregate stages of retention over time (e.g. 6, 12, and 18 months) or add stages for treatment milestones unique to a specific service setting (7). For example, Khalid et al.’s (14) OUD Cascade model for office-based buprenorphine treatment considers the referral for treatment (Stage 1), scheduling of an initial visit (Stage 2) and completion of an initial visit (Stage 3) as three distinct stages.

This paper describes OUD Cascade of Care implementation at New York City (NYC) Health+Hospitals (“H+H”). H+H is the largest urban public hospital system in the United States, annually encountering over one million unique patients, about 128,000 with substance use disorders (15). We describe operationalization of the H+H OUD Cascade of Care (hereafter “the H+H Cascade” or “the Cascade”), including decisions for compiling data/defining measures, and challenges/solutions. This case example will inform other hospitals and health systems in developing their own Cascade frameworks and applying the Cascade to promote uptake of evidence-based practices to improve health outcomes and reduce mortality for patients with OUD.

## METHODS

### Setting

H+H comprises over 70 locations across NYC, including eleven acute care hospitals, five post-acute/long-term care facilities, and 56 Federally Qualified Health Center (FQHC) clinics called Gotham Health (16). H+H is a safety net institution, serving patients regardless of documentation, insurance status or ability to pay (15). H+H’s Office of Behavioral Health (“OBH”), provides system-wide leadership, and support for all behavioral health services. Given its mandate to combat the opioid epidemic, OBH operationalized an OUD Cascade of Care quality improvement tool to identify optimal resource deployment. Members of OBH’s data (EC, JG) and leadership (NI, DS) teams met iteratively with OUD Cascade researchers (ARW, NK, BH) to design the “H+H Cascade” (define stages and outcome measures) using H+H EHR data.

### Compiling Patient Data

The OBH data team obtained and synthesized historical patient data between 2017 and 2021 from the following EHR and electronic billing systems used across H+H since 2017: EPIC (EHR), Quadramed (EHR), Soarian (billing), and Unity (billing). In 2013, H+H began condensing these systems to improve efficiency, first transitioning EHR to EPIC and billing to Soarian, then from 2017-2019, moving exclusively to EPIC for EHR and billing (16). H+H’s Office of Population Health, which oversees research and evaluation, linked unique patient records across all systems, storing linked encounter and billing records in a Microsoft Structured Query Language (SQL) Server (a standard programming language for accessing data in a relational database) (17). To join patient records across systems, a unique patient ID was created from concatenated identifiers within each system (name, sex, and date of birth).

The OBH data team used the SQL Data Warehouse to construct the H+H Cascade from patient/encounter level data containing dates of service, service location (facility; department), primary and non-primary billing diagnoses, and patient demographics. Data sets were exported directly from EPIC, using pre-built reports documenting patient encounters, medications, and diagnoses from the Problem List. Historical medication administration and prescription data were exported from Quadramed.

## RESULTS

### Cascade Measurement Design and Scope

We designed the H+H Cascade to be a single population, longitudinal, and denominator-denominator linked analysis, following the same cohort of patients over time. Patients must have reached the prior stage to progress to subsequent stages. We included patients over age 18 with at least one inpatient, outpatient, or emergency department encounter at any H+H facility.

*Scope*: We defined 5 stages ranging from OUD diagnosis to MOUD 6-month retention (Figure 1). Given a lack of “true” population-based estimates of OUD in our target population, we defined Stage 1 as any patient having a documented OUD diagnosis in patient EHR or billing records. Our final stage represents patient achievement of 6-month retention on MOUD as the EHR could not systematically reflect full sustained remission of OUD, a common limitation to EHR systems (18). Note that Figure 1 presents hypothetical data assuming a 33% drop off from stage to stage as a rough proxy for attrition across progressive Cascade stages (10). Detailed operational definitions for each stage follow in the next section.

1. Diagnosed with OUD
2. Engaged with addiction services
3. Initiated MOUD treatment in an outpatient setting
4. Had MOUD follow up visit within 34 days of initiation
5. Retained on MOUD for 6+ months

**Figure 1:**
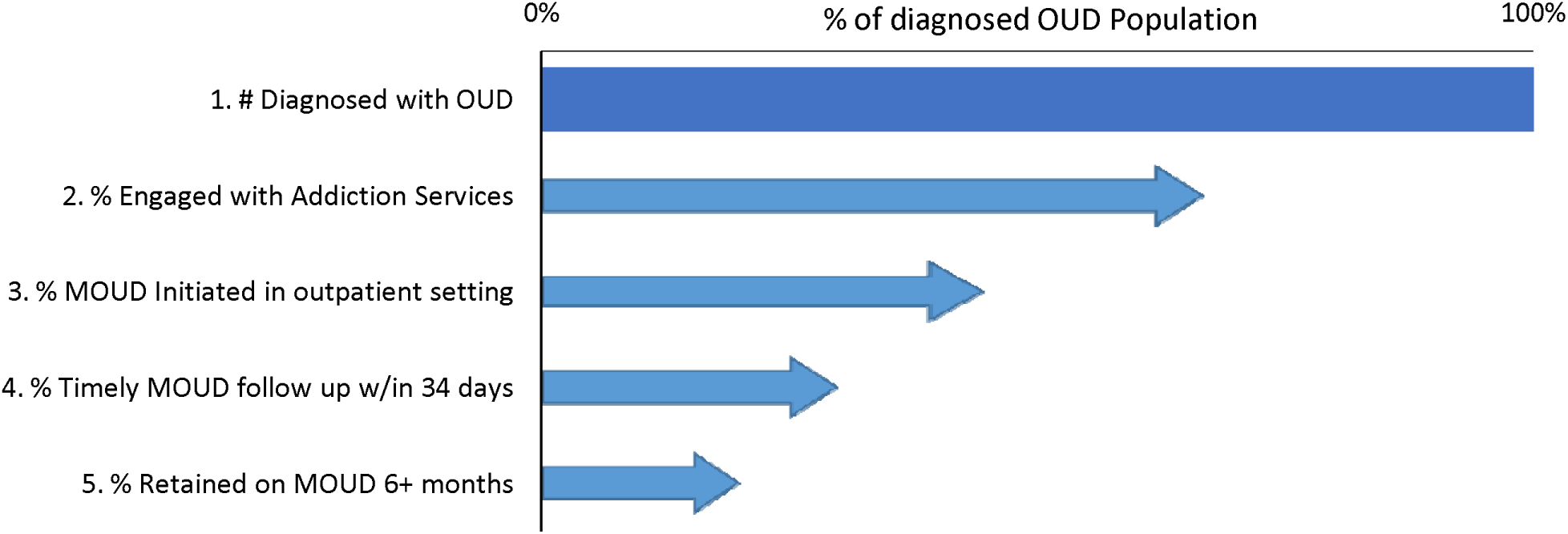
OUD Cascade of Care Stage Design through 6-Month Retention (Theoretical) Notes: We defined 5 stages ranging from OUD diagnosis to MOUD 6-month retention. We defined Stage 1 as any patient having a documented OUD diagnosis in patient EHR or billing records. Our final stage represents patient achievement of 6-month retention on MOUD as the EHR could not systematically reflect full sustained remission of OUD, a common limitation to EHR systems (18). Note that the Figure presents hypothetical data assuming a 33% drop off from stage to stage as a proxy for attrition across progressive Cascade stages. Detailed operational definitions for each stage are in the text. Lengths of bars do not represent actual numbers and are hypothetical. OUD=opioid use disorder. MOUD=medications for opioid use disorder.

### Operational Definitions for Stages of the Cascade

Criteria for inclusion and data sources for each Cascade stage are outlined in Table 1 and further described in the following sections.

**Table 1:** Inclusion Criteria and Data Sources for the H+H OUD Cascade of Care.

| Stage | Inclusion Criteria | Data Source/s |
| --- | --- | --- |
| 1. Diagnosed OUD | Patients with either an inpatient, outpatient, or emergency encounter at any H+H facility with an ICD-10 diagnosis of OUD documented as the primary or non-primary billing diagnosis; or patients with an OUD diagnosis code documented as an active problem on their EPIC Problem List. See Appendix for list of codes. | EPIC and Quadramed encounter and billing data stored in SQL Data Warehouse; EPIC Problem List report |
| 2. Engaged with addiction services | Patients that had an inpatient, emergency dept, or outpatient encounter in one of four possible addiction treatment settings. This could be the same encounter as their diagnostic encounter, or a separate encounter on or after the diagnostic encounter. See Table 2 for details on included addiction treatment settings. | EPIC and Quadramed encounter data stored in SQL Data Warehouse; EPIC Program-Specific Reports |
| 3. Initiated MOUD treatment in an outpatient setting | Patients with either 1+ outpatient prescriptions for buprenorphine or IM naltrexone; or patients that had a methadone encounter in an OTP. The date of the prescription or the OTP encounter could occur on the same date/encounter or after the date/encounter they engaged with addiction services. | EPIC and Quadramed encounter data stored in SQL Data Warehouse; EPIC Medication Reports; Archived Quadramed Medication Reports |
| 4. Had a timely MOUD follow up visit within 34 days of initiation | Patients with either 1+ subsequent outpatient prescription for buprenorphine or IM naltrexone within 34 days of the prior prescription; or patients that had 1+ subsequent methadone encounters in an OTP within 34 days of their prior encounter. | EPIC and Quadramed encounter data stored in SQL Data Warehouse; EPIC Medication Reports; Archived Quadramed Medication Reports |
| 5. Retained on MOUD for 6 months or more | Patients with subsequent outpatient prescriptions for buprenorphine or naltrexone 6 months after initiation; or patients that had subsequent methadone encounters in an OTP 6 months after initiation. Within the six month retention period, patients with a gap in prescription coverage longer than 30 days or patients who did not return to an OTP within 30 days will be excluded. | EPIC and Quadramed encounter data stored in SQL Data Warehouse; EPIC Medication Reports; Archived Quadramed Medication Reports |

**Notes:**
OUD = Opioid Use Disorder
H+H = NYC Health + Hospitals
MOUD = Medications for Opioid Use Disorder
IM naltrexone = intramuscular naltrexone
OTP = Opioid Treatment Program
SQL = Structured Query Language
EPIC and Quadramed are two electronic health record systems.

#### Stage 1: Diagnosed with Opioid Use Disorder (OUD)

Patients were included in Stage 1 if they had at least one inpatient, outpatient, or emergency department encounter at any H+H facility with an ICD-10 diagnosis of OUD documented as the primary or non-primary billing diagnosis in EPIC or Quadramed. ICD-10 codes included are listed in the Appendix. Patients were also included in Stage 1 if they had an OUD diagnosis documented as an active problem on their EPIC Problem List. The EPIC Problem List in each patient’s chart allows providers to document current/past diagnoses. To capture patients with new (index) OUD diagnoses and exclude patients with prior diagnoses who might already be active in treatment, we established a lookback period six months prior to the data observation period. If patients had a documented OUD diagnosis in the past 6 months, they were excluded.

#### Stage 2: Engaged with addiction services

Patients were included in Stage 2 if they contacted or were reached by addiction specialty services at H+H following their diagnosis. Treatment engagement could consist of one or more completed encounters occurring across four treatment settings throughout the system. We counted engagement whether it occurred with the same encounter as OUD diagnosis, or after the diagnostic encounter. Given heterogeneity across settings, each service setting or intervention had a unique set of inclusion criteria, delineated in Table 2.

**Table 2:** Engagement with addiction services across health system settings.

| **Treatment Setting** | **Description** | **Criteria to meet OUD Cascade Stage 2 Engagement** |
| --- | --- | --- |
| Specialty Substance Use Disorder Clinics | H+H operates 10 NY state-licensed outpatient chemical dependency clinics and four outpatient Opioid Treatment Programs (OTPs). | If a patient completed a visit in any of these clinics, they were included in this stage. No particular diagnosis was required for inclusion as these are addiction specialty treatment settings. |
| Outpatient Healthcare Settings (Primary Care; Infectious Disease Clinics) | H+H offers Office-Based Opioid Treatment (OBOT) in some primary care settings. Additionally, patients who receive treatment for HIV or HCV may receive treatment for OUD in specialty outpatient departments at H+H, specifically Virology and Infectious Disease clinics. | As available data sets did not distinguish substance use treatment encounters from other care encounters in these settings, patients were considered engaged in treatment if OUD was documented as the primary billing diagnosis for the completed visit. |
| Emergency Department (ED) Settings | ED settings encompassed both adult general medical EDs and specialty psychiatry ED settings. ED settings at H+H have providers that can prescribe MOUD, as well as “ED Leads”, an addiction service located in the ED that consists of social workers and peers that provide education, SBIRT* and follow up such as peer support services. | Patients seen in the ED were included if either:<br>1. OUD was documented as the primary billing diagnosis for the visit and they were administered buprenorphine or methadone during the visit.<br>2. Patients had an encounter with an ED Leads team member, regardless of OUD diagnosis |
| Inpatient Settings | CATCH (Consult for Addiction Treatment and Care in Hospitals), is an inpatient consult service consisting of prescribers, social workers, and peers that is located at six of the eleven acute care facilities at H+H. In addition to inpatient consultation, CATCH teams provide wraparound care in the outpatient setting by providing bridging services to ongoing maintenance programs (37). | Patients met criteria for the engagement stage if they had any documented encounter with a CATCH team member, regardless of OUD diagnosis. |

\* Screening, Brief Intervention, and Referral to Treatment
**Notes:**
OUD = Opioid Use Disorder
H+H = NYC Health + Hospitals
HIV = Human Immunodeficiency Virus
HCV = Hepatitis C
MOUD = Medications for Opioid Use Disorder
IM naltrexone = intramuscular naltrexone

#### Stage 3: Initiated MOUD treatment in an outpatient setting

Patients were included in Stage 3 if they initiated MOUD in tandem with or following their engagement in addiction services at an outpatient setting providing MOUD maintenance. Receipt of MOUD in inpatient or ED settings (Stage 2) did not qualify because this stage aims to capture initiation of maintenance medication therapy, not receipt of medications given to treat withdrawal. Medications included in this stage were: buprenorphine (all formulations except those indicated solely for pain conditions), methadone, and intramuscular extended release (“xr-“) naltrexone. A list of included formulations is provided in the Appendix. Medication data for buprenorphine and xr-naltrexone was accessible through the EHR as documented in medication orders. Methadone administrations are recorded in a software system that is not integrated with the patient’s EHR; therefore, we counted a completed Outpatient Treatment Program (“OTP”) visit as a proxy for methadone administration, thereby excluding use of methadone for pain.

This stage was adapted from the National Quality Forum (NQF) measure for the use of pharmacotherapy for OUD (NQF 3400) (19). Although counseling and behavioral therapies can support patients as part of their treatment, MOUD is considered the gold standard for treating OUD (20). However, fewer than one third of patients in substance use treatment programs for OUD receive medication (21).

#### Stage 4: MOUD follow up visit within 34 days of initiation

Patients were included in Stage 4 if they had a timely MOUD follow up after their MOUD initiation in an outpatient setting, which was adapted from the Healthcare Effectiveness Data and Information Set (HEDIS) (22, 23) addiction treatment engagement quality measure (24). Patients had to have at least one subsequent medication order for buprenorphine or xr-naltrexone or documented visit in an OTP within 34 days of initiation. Patients with no prescriptions or OTP visits within 30 days from MOUD initiation were excluded and considered discontinued in care.

Due to data limitations, we established rules to determine length of medication coverage to assess MOUD engagement and retention. EPIC data typically contained a start and end date for a prescription, but Quadramed data only contained prescription start dates. Therefore, we created a coverage rule for buprenorphine prescriptions without end dates, where we imputed an end date of 30 days following the start date, consistent with common clinical practice of prescribing no more than a 30 day supply (25). Xr-naltrexone was also considered to cover a patient for 30 days given it is a monthly injection. For OTP visits for which we did not have access to methadone dispensing data or data on take home doses (OTPs manage care through a separate EHR), we established a rule that one visit in an OTP would “cover” a patient for 3 days, in line with federal guidelines around methadone dosing for new patients (25). Patients who had gaps in care of longer than 30 days between prescriptions or OTP visits were considered discontinued.

#### Stage 5: Retained on MOUD for 6+ months

Patients were included in Stage 5 if they were retained on MOUD for at least six months, consistent with the NQF-endorsed quality measure for continuity of pharmacotherapy of MOUD for a minimum of 180 days (NQF #3175) (26). Patients that had medication orders (buprenorphine or xr-naltrexone), or OTP visits 6 months after their initial MOUD prescription or visit were included in this stage, as long as they did not have a gap between prescriptions or visits longer than 30 days, following the coverage assumption rules defined above. NQF measure #3175 defines the gap at the 7 day mark (26), but our team chose to extend the gap to 30 days to account for patients who were unable to return to the hospital for a follow up prescription appointment within that small of a time window, which may have been an issue during the early days of the COVID-19 pandemic.

When fully deploying the Cascade, we are considering testing dynamic definitions of follow-up engagement (Stage 4) and retention (Stage 5) via sensitivity analyses using different cutoff periods to determine treatment discontinuation. Although we operationalized our allowable treatment gap at 30 days, the existing literature on medication gaps ranges between 5 to 90 days (27). In a study measuring the duration of buprenorphine treatment episodes, Dong et al. (27) demonstrated that changing the allowable gap between MOUD episodes (their team analyzed gaps at 7, 14, 30, and 60 days) impacted the researchers’ understanding of patients’ continuity of care. This is particularly relevant as federal regulations for OTP visits and methadone take-home doses have changed in response to COVID-19. Prior to the pandemic, patients starting treatment up until their first 90 days in care were required to visit an OTP daily and patients in the second 90 days of treatment could visit every other day and were allowed up to two weekly take-home doses (25). In March 2020, the Substance Abuse and Mental Health Services Administration (SAMHSA) relaxed regulations, issuing a waiver permitting up to 28 days of take homes for stable patients and up to 14 days of take homes for less stable patients (28). We anticipate that these changes will impact the rules we establish regarding medication coverage.

Summary statistics are presented in Table 3 showing the total number of individuals identified with OUD in the H+H system (n= 33,616) and their progression through sequential stages of the Cascade. Consistent with prior studies (14), we found that attrition was most steep in earlier stages of the Cascade.

**Table 3:** OUD Cascade of Care in a Large Public Health System.

|  | 1. # Diagnosed with OUD |  | 2. Engaged with Addiction Services |  | 3. MOUD Initiated in outpatient setting |  | 4. Timely MOUD follow up w/in 34 days |  | 5. Retained on MOUD 6+ months |  |
| --- | --- | --- | --- | --- | --- | --- | --- | --- | --- | --- |
|  | N = 33616 |  | N = 16203 |  | N = 3587 |  | N = 2281 |  | N = 1196 |  |
| **Age** |  |  |  |  |  |  |  |  |  |  |
| 18-25 | 2062 | 6.13% | 933 | 5.76% | 186 | 5.19% | 101 | 4.43% | 38 | 3.18% |
| 26-35 | 7103 | 21.13% | 3558 | 21.96% | 880 | 24.53% | 556 | 24.38% | 261 | 21.82% |
| 36-45 | 6822 | 20.29% | 3455 | 21.32% | 793 | 22.11% | 511 | 22.40% | 262 | 21.91% |
| 46-55 | 8173 | 24.31% | 4077 | 25.16% | 880 | 24.53% | 574 | 25.16% | 306 | 25.59% |
| 56-65 | 6896 | 20.51% | 3224 | 19.90% | 673 | 18.76% | 433 | 18.98% | 263 | 21.99% |
| 66 and up | 2505 | 7.45% | 937 | 5.78% | 175 | 4.88% | 106 | 4.65% | 66 | 5.52% |
| Unknown | 55 | 0.16% | 19 | 0.12% | 0 | 0.00% | 0 | 0.00% | 0 | 0.00% |
| **Sex** |  |  |  |  |  |  |  |  |  |  |
| Female | 8798 | 26.17% | 3632 | 22.42% | 815 | 22.72% | 506 | 22.18% | 270 | 22.58% |
| Male | 24815 | 73.82% | 12568 | 77.57% | 2770 | 77.22% | 1775 | 77.82% | 926 | 77.42% |
| Unknown | 3 | 0.01% | 3 | 0.02% | 2 | 0.06% | 0 | 0.00% | 0 | 0.00% |
| **Race** |  |  |  |  |  |  |  |  |  |  |
| Asian | 502 | 1.49% | 249 | 1.54% | 71 | 1.98% | 50 | 2.19% | 29 | 2.42% |
| Black | 11041 | 32.84% | 5401 | 33.33% | 905 | 25.23% | 554 | 24.29% | 263 | 21.99% |
| Hispanic/Latinx | 4367 | 12.99% | 2186 | 13.49% | 417 | 11.63% | 256 | 11.22% | 138 | 11.54% |
| Native American or Alaskan Native | 100 | 0.30% | 64 | 0.39% | 16 | 0.45% | 9 | 0.39% | 6 | 0.50% |
| Native Hawaiian or Other Pacific Islander | 16 | 0.05% | 5 | 0.03% | 3 | 0.08% | 2 | 0.09% | 0 | 0.00% |
| Other | 10407 | 30.96% | 4603 | 28.41% | 1095 | 30.53% | 693 | 30.38% | 365 | 30.52% |
| White | 6728 | 20.01% | 3537 | 21.83% | 1046 | 29.16% | 703 | 30.82% | 390 | 32.61% |
| Unknown | 455 | 1.35% | 158 | 0.98% | 34 | 0.95% | 14 | 0.61% | 5 | 0.42% |

## DISCUSSION

### Preliminary Implementation of the H+H Cascade

Prior to the Cascade’s creation, H+H leadership tracked the overall number of patients entering our system with diagnosed OUD, and how many received MOUD, but it was unclear how many patients were connected to interventions at various points within the system or where they were lost to follow up. The strength of the Cascade framework is its ability to help the system identify trends and gaps in care receipt and where to target clinical interventions to ensure that a larger percentage of patients receives evidence-based services. Using preliminary datasets in the Cascade model, the H+H OBH team developed an internal dashboard that displays aggregate patient data for each cascade stage. The dashboard also shows how many patients and in which treatment setting(s) they engaged in treatment (Stage 2) following diagnosis (see Table 2 for all included treatment settings). Preliminary analyses have shown that the highest volume of patients engage in care through H+H’s “ED Leads” addiction teams, which consist of social workers and peers that provide education, SBIRT, and follow up services within the Emergency Departments. By highlighting this program as an important intervention to connect OUD patients to continuing treatment, OBH leadership has been able to advocate for an increase in resources and staffing to enhance the program.

The clarity and consistency of demonstrating the system’s reach of patients through the Cascade stages can be easily translated to clinical staff, hospital executives, researchers, and other stakeholders. The model highlights areas ripe for improvement in an easily digestible way while also having the power to disaggregate complex data when more information is needed about where the system is and is not effectively intervening with patients. H+H’s internal dashboard displays patient data from the entire system that can be filtered by hospital location. The ability to toggle between system-wide and hospital-specific views of the Cascade can help clarify system vs. site-specific needs, and the model has already been applied to advocate for quality improvement interventions at H+H. A preliminary comparative analysis of a system-wide and a hospital-specific Cascade showed that a lower rate of patients had initiated MOUD (Stage 3) at one hospital in comparison to the rate for the entire system. The OBH team used this analysis to alert hospital leadership to the discrepancy, and advocated for an intervention that would better connect patients to medication.

### Challenges and Solutions

#### Data Access and Linkages

The experience of H+H shows the feasibility of building an OUD Cascade of Care in a large health system even with shifts in data management software. We show that committing to stage definitions with fidelity is only possible after understanding a system’s data constraints and responding to challenges accordingly.

Although our team had access to a wide variety of data sources, the complexity of H+H’s data systems limited our ability to access certain records and understand linkages over time. Because we relied on EPIC and Quadramed datasets, we were unable to rely on a numerical identifier attached to the patient in both EHR systems. To work around this, we developed our own unique patient identifier that concatenated the first 5 digits of a patient’s last name, the first 2 digits of a patient’s first name, an 8 digit birthdate, and an F or M initial for the patient’s sex. This process was undertaken using SQL queries and required several rounds of manual review and validation checks. Patient name misspellings, changes in sex, or other data inconsistences may have contributed to one patient having more than one identifier, despite our best efforts to develop a unique ID for each person.

While compiling and synthesizing data sources across a large hospital system allows us to capture patients who may have received treatment in multiple H+H facilities, our analysis is limited by our inability to access data outside of the system. There may be patients with OUD included in our sample who sought treatment or initiated/discontinued MOUD at another facility outside of the H+H system but could not be identified. Additionally, because our research team did not have access to insurance claims data, part of the criteria for medication initiation and subsequent stages relied on prescription order records, which could result in the assumption that a patient adhered to their medication when they did not in fact fill their prescription or take the medication. Thus, analyses that can track multi-system service utilization using claims data are also needed to understand true patient trajectories.

Data disaggregation using patient demographics is also a critical step in identifying population level disparities in access to care. Given the significant racial and ethnic disparities in receipt of OUD services (29), the Cascade can be an important tool in understanding where a healthcare system is best engaging certain patient groups, and where additional organizational level strategies are needed to deliver culturally competent care. The encounter-level datasets compiled for the Cascade contain fields that indicate patient race, ethnicity, and sex, therefore it is possible to investigate potential discrepancies in access and retention in care. However, due to issues including limited race and ethnicity category options within the EHR and patients’ unwillingness to report, some data may be missing or a race/ethnicity category may be unspecified. The ability to link datasets by patient identifier allows for the reduction of some missing data. If demographic data is missing from one patient record, but a patient has returned for an encounter within the system and demographic data is entered, the records can be linked to substitute for the missing data. Steps will be taken in future analyses to reduce missing and unspecified variables.

#### Defining Measures

The accurate measurement of the overall patient population may be impacted by the use of diagnosis codes as the main indicator to identify patients with OUD. Stigma may prevent patients who use opioids from disclosing use to healthcare providers or seeking treatment (30, 31). This may contribute to an undercount of patients analyzed using the model. Other models may consider adding an “at risk” stage to analyze patients who may not meet the clinical criteria for OUD but take opioids for pain management or score within a certain range on validated substance use screeners including the DAST-10 or CAGE-AID (10, 32, 33, 34).

Another ongoing challenge is establishing rules for defining gaps in care and patient drop offs between stages. We established a rule for MOUD engagement and retention that if a patient had a coverage gap of longer than 30 days, they would not achieve the subsequent retention stage. Creating rules around patient drop off due to a coverage gap is complex given the nature of OUD as patients may start and stop MOUD multiple times (35, 36). While establishing an exclusion rule may mean that fewer patients can achieve subsequent stages, we chose to initially design the Cascade based on established standards (e.g. HEDIS or NQF-endorsed measures) as a quality improvement tool for the H+H system.

### Conclusion and Next Steps

Despite data and measurement challenges, our experience designing the H+H Cascade demonstrates the feasibility of other large healthcare systems creating their own models. While H+H’s stages were adapted from national quality assurance measures for MOUD initiation, timely follow up, and retention, that have fixed definitions, the Cascade model is flexible enough for systems to design their own models with inclusion rules based off of the programs and services they offer. For example, as shown in Table 2, patients progressed to Stage 2 if they engaged in services in at least one of four addiction treatment settings. These settings included programs and interventions offered at H+H, but other systems could design criteria that would reflect their unique services. Similarly, another system may want to expand the Cascade’s depth and define monthly stages for medication retention to investigate patient dropoff at smaller intervals before the NQF defined 6 month mark. Another system may choose to change retention stage criteria to require completion of a follow up visit in a specific setting, in addition to MOUD receipt.

Now that the H+H Cascade has been established, the OBH data and leadership teams will use the framework to quantify and evaluate H+H patient progression through each stage of treatment. Immediate next steps include determining a timeframe for and undergoing an analysis of the H+H patient population within the Cascade framework.

We plan on using that analysis to understand OUD prevalence in the system, identify gaps in treatment, and examine predictors and risk factors for continuation or drop off. Consistent with prior studies, we found that drop-offs in care disproportionately occur in earlier stages (14), which has implications for system-wide promotion and implementation of intensive services in response to new diagnoses and early engagement. Subsequently, building on our preliminary internal dashboard, we aim to create and validate a data model to apply the framework in real time and assess its utility as a tool to support policy innovation and programmatic change to direct H+H resources towards strategic areas of intervention and ultimately improve patient outcomes.

Through the creation of more robust centralized surveillance tools such as the OUD Cascade described here, H+H and other large health systems can be more data-driven and efficient in how they address OUD and overdose. Because Cascade milestones carefully align with SUD-related HEDIS/NQF measures, not only will the Cascade help move the needle on the system’s quality improvement efforts, but will also be useful for reducing costs, without creating redundant reporting requirements.

Expanding these surveillance efforts can facilitate access to evidence-based treatment and engagement of high risk/complex patients at critical points of intervention with health systems to address the overdose crisis and improve care for a historically marginalized population.

## Data Availability

Aggregated data are presented in the manuscript

## Notes

### Competing Interest Statement

Dr. Williams receives consulting fees, equity, and travel reimbursement from Ophelia Health Inc a telehealth company for the treatment of opioid use disorder. He also receives consulting fees from the National Quality Forum. Dr. Krawczyk receives fees for expert testimony in ongoing opioid litigation.
Dr. Krawczyk was supported by the National Institute On Drug Abuse of the National Institutes of Health under Award Number K01DA055758. The content is solely the responsibility of the authors and does not necessarily represent the official views of the National Institutes of Health.
The other authors report no relevant disclosures.

### Funding Statement

Dr. Krawczyk was supported by the National Institute On Drug Abuse of the National Institutes of Health under Award Number K01DA055758. The content is solely the responsibility of the authors and does not necessarily represent the official views of the National Institutes of Health.

### Author Declarations

The study was approved by the BRANY Institutional Review Board.

