## Appendix for "Application of an Opioid Use Disorder Cascade of Care in a Large Public Health System"

| **OUD Diagnosis Codes** | |
| --- | --- |
| **ICD-10-CM Code** | **ICD-10-CM Description** |
| F11.10 | Opioid abuse, uncomplicated |
| F11.11 | Opioid abuse, in remission |
| F11.120 | Opioid abuse with intoxication, uncomplicated |
| F11.121 | Opioid abuse with intoxication delirium |
| F11.122 | Opioid abuse with intoxication with perceptual disturbance |
| F11.129 | Opioid abuse with intoxication, unspecified |
| F11.13 | Opioid abuse with withdrawal |
| F11.14 | Opioid abuse with opioid-induced mood disorder |
| F11.150 | Opioid abuse with opioid-induced psychotic disorder with delusions |
| F11.151 | Opioid abuse with opioid-induced psychotic disorder with hallucinations |
| F11.159 | Opioid abuse with opioid-induced psychotic disorder, unspecified |
| F11.181 | Opioid abuse with opioid-induced sexual dysfunction |
| F11.182 | Opioid abuse with opioid-induced sleep disorder |
| F11.188 | Opioid abuse with other opioid-induced disorder |
| F11.19 | Opioid abuse with unspecified opioid-induced disorder |
| F11.20 | Opioid dependence, uncomplicated |
| F11.21 | Opioid dependence, in remission |
| F11.220 | Opioid dependence with intoxication, uncomplicated |
| F11.221 | Opioid dependence with intoxication delirium |
| F11.222 | Opioid dependence with intoxication with perceptual disturbance |
| F11.229 | Opioid dependence with intoxication, unspecified |
| F11.23 | Opioid dependence with withdrawal |
| F11.24 | Opioid dependence with opioid-induced mood disorder |
| F11.250 | Opioid dependence with opioid-induced psychotic disorder with delusions |
| F11.251 | Opioid dependence with opioid-induced psychotic disorder with hallucinations |
| F11.259 | Opioid dependence with opioid-induced psychotic disorder, unspecified |
| F11.281 | Opioid dependence with opioid-induced sexual dysfunction |
| F11.282 | Opioid dependence with opioid-induced sleep disorder |
| F11.288 | Opioid dependence with other opioid-induced disorder |
| F11.29 | Opioid dependence with unspecified opioid-induced disorder |
| F11.90 | Opioid use, unspecified, uncomplicated |
| F11.920 | Opioid use, unspecified, with intoxication, uncomplicated |
| F11.921 | Opioid use, unspecified, with intoxication delirium |
| F11.922 | Opioid use, unspecified, with intoxication with perceptual disturbance |
| F11.929 | Opioid use, unspecified, with intoxication, unspecified |
| F11.93 | Opioid use, unspecified with withdrawal |
| F11.94 | Opioid use, unspecified with opioid-induced mood disorder |
| F11.950 | Opioid use, unspecified with opioid-induced psychotic disorder with delusions |
| F11.951 | Opioid use, unspecified with opioid-induced psychotic disorder with hallucinations |
| F11.959 | Opioid use, unspecified with opioid-induced psychotic disorder, unspecified |
| F11.981 | Opioid use, unspecified with opioid-induced sexual dysfunction |
| F11.982 | Opioid use, unspecified with opioid-induced sleep disorder |
| F11.988 | Opioid use, unspecified with other opioid-induced |
| F11.99 | Opioid use, unspecified with unspecified opioid-induced disorder |
| T40.0X1 | Included miscode |
| T40.0X1A | Poisoning by opium, accidental (unintentional), initial encounter |
| T40.0X1D | Poisoning by opium, accidental (unintentional), subsequent encounter |
| T40.0X1S | Poisoning by opium, accidental (unintentional), sequela |
| T40.0X2 | Included miscode |
| T40.0X2A | Poisoning by opium, intentional self-harm, initial encounter |
| T40.0X2D | Poisoning by opium, intentional self-harm, subsequent encounter |
| T40.0X2S | Poisoning by opium, intentional l self-harm, sequela |
| T40.0X3 | Included miscode |
| T40.0X3A | Poisoning by opium, assault, initial encounter |
| T40.0X3D | Poisoning by opium, assault subsequent encounter |
| T40.0X3S | Poisoning by opium, , assault, sequela |
| T40.0X4 | Included miscode |
| T40.0X4A | Poisoning by opium, undetermined, initial encounter |
| T40.0X4D | Poisoning by opium, undetermined, subsequent encounter |
| T40.0X4S | Poisoning by opium, undetermined, sequela |
| T40.0X5 | Included miscode |
| T40.0X5A | Adverse effect of opium, initial encounter |
| T40.0X5D | Adverse effect of opium, subsequent encounter |
| T40.0X5S | Adverse effect of opium, sequela |
| T40.1X1 | Included miscode |
| T40.1X1A | Poisoning by heroin, accidental (unintentional), initial encounter |
| T40.1X1D | Poisoning by heroin, accidental (unintentional), subsequent encounter |
| T40.1X1S | Poisoning by heroin, accidental (unintentional), sequela |
| T40.1X2 | Included miscode |
| T40.1X2A | Poisoning by heroin, intentional self-harm, initial encounter |
| T40.1X2D | Poisoning by heroin, intentional self-harm, subsequent encounter |
| T40.1X2S | Poisoning by heroin, intentional self-harm, sequela |
| T40.1X3 | Included miscode |
| T40.1X3A | Poisoning by heroin, assault, initial encounter |
| T40.1X3D | Poisoning by heroin, assault, subsequent encounter |
| T40.1X3S | Poisoning by heroin, assault, sequela |
| T40.1X4 | Included miscode |
| T40.1X4A | Poisoning by heroin, undetermined, initial encounter |
| T40.1X4D | Poisoning by heroin, undetermined, subsequent encounter |
| T40.1X4S | Poisoning by heroin, undetermined, sequela |
| T40.1X5 | Included miscode |
| T40.1X5A | Adverse effect of heroin initial encounter |
| T40.1X5D | Adverse effect of heroin subsequent encounter |
| T40.1X5S | Adverse effect of heroin sequela |
| T40.2X1 | Included miscode |
| T40.2X1A | Poisoning by other opioids, accidental (unintentional), initial encounter |
| T40.2X1D | Poisoning by other opioids, accidental (unintentional), subsequent encounter |
| T40.2X1S | Poisoning by other opioids, accidental (unintentional), sequela |
| T40.2X2 | Included miscode |
| T40.2X2A | Poisoning by other opioids, intentional self-harm, initial encounter |
| T40.2X2D | Poisoning by other opioids, intentional self-harm, subsequent encounter |
| T40.2X2S | Poisoning by other opioids, intentional self-harm, sequela |
| T40.2X3 | Included miscode |
| T40.2X3A | Poisoning by other opioids, assault, initial encounter |
| T40.2X3D | Poisoning by other opioids, assault, subsequent encounter |
| T40.2X3S | Poisoning by other opioids, assault, sequela |
| T40.2X4 | Included miscode |
| T40.2X4A | Poisoning by other opioids, undetermined, initial encounter |
| T40.2X4D | Poisoning by other opioids, undetermined, subsequent encounter |
| T40.2X4S | Poisoning by other opioids, undetermined, sequela |
| T40.2X5 | Included miscode |
| T40.2X5A | Adverse effect of other opioids, initial encounter |
| T40.2X5D | Adverse effect of other opioids, subsequent encounter |
| T40.2X5S | Adverse effect of other opioids, sequela |
| T40.3X1 | Included miscode |
| T40.3X1A | Poisoning by methadone, accidental (unintentional), initial encounter |
| T40.3X1D | Poisoning by methadone, accidental (unintentional), subsequent encounter |
| T40.3X1S | Poisoning by methadone, accidental (unintentional), sequela |
| T40.3X2 | Included miscode |
| T40.3X2A | Poisoning by methadone, intentional self-harm, initial encounter |
| T40.3X2D | Poisoning by methadone, intentional self-harm, subsequent encounter |
| T40.3X2S | Poisoning by methadone, intentional self-harm, sequela encounter |
| T40.3X3 | Included miscode |
| T40.3X3A | Poisoning by methadone, assault, initial encounter |
| T40.3X3D | Poisoning by methadone, assault, subsequent encounter |
| T40.3X3S | Poisoning by methadone, assault, sequela encounter |
| T40.3X4 | Included miscode |
| T40.3X4A | Poisoning by methadone, undetermined, initial encounter |
| T40.3X4D | Poisoning by methadone, undetermined, subsequent encounter |
| T40.3X4S | Poisoning by methadone, undetermined, sequela |
| T40.3X5 | Included miscode |
| T40.3X5A | Adverse effect of methadone, initial encounter |
| T40.3X5D | Adverse effect of methadone, subsequent encounter |
| T40.3X5S | Adverse effect of methadone, sequela |
| T40.4X1 | Included miscode |
| T40.4X1A | Poisoning by synthetic narcotics, accidental (unintentional), initial encounter |
| T40.4X1D | Poisoning by synthetic narcotics, accidental (unintentional), subsequent encounter |
| T40.4X1S | Poisoning by synthetic narcotics, accidental (unintentional), sequela |
| T40.4X2 | Included miscode |
| T40.4X2A | Poisoning by other synthetic narcotics, intentional self-harm, initial encounter |
| T40.4X2D | Poisoning by other synthetic narcotics, intentional self-harm, subsequent encounter |
| T40.4X2S | Poisoning by other synthetic narcotics, intentional self-harm, sequela |
| T40.4X3 | Included miscode |
| T40.4X3A | Poisoning by other synthetic narcotics, assault, initial encounter |
| T40.4X3D | Poisoning by other synthetic narcotics, assault, subsequent encounter |
| T40.4X3S | Poisoning by other synthetic narcotics, assault, sequela |
| T40.4X4 | Included miscode |
| T40.4X4A | Poisoning by synthetic narcotics, undetermined, initial encounter |
| T40.4X4D | Poisoning by synthetic narcotics, undetermined, subsequent encounter |
| T40.4X4S | Poisoning by synthetic narcotics, undetermined, sequela |
| T40.4X5 | Included miscode |
| T40.4X5A | Adverse effect of synthetic narcotics, initial encounter |
| T40.4X5D | Adverse effect of synthetic narcotic, subsequent encounter |
| T40.4X5S | Adverse effect of synthetic narcotic, sequela |
| T40.601 | Included miscode |
| T40.601A | Poisoning by unspecified narcotics, accidental (unintentional), initial encounter |
| T40.601D | Poisoning by unspecified narcotics, accidental (unintentional), subsequent encounter |
| T40.601S | Poisoning by unspecified narcotics, accidental (unintentional), sequela |
| T40.602 | Included miscode |
| T40.602A | Poisoning by unspecified narcotics, intentional self-harm, initial encounter |
| T40.602D | Poisoning by unspecified narcotics, intentional self-harm, subsequent encounter |
| T40.602S | Poisoning by unspecified narcotics, intentional self-harm, sequela encounter |
| T40.603 | Included miscode |
| T40.603A | Poisoning by unspecified narcotics, assault, initial encounter |
| T40.603D | Poisoning by unspecified narcotics, assault, subsequent encounter |
| T40.603S | Poisoning by unspecified narcotics, assault, sequela |
| T40.604 | Included miscode |
| T40.604A | Poisoning by unspecified narcotics, undetermined, initial encounter |
| T40.604D | Poisoning by unspecified narcotics, undetermined, subsequent encounter |
| T40.604S | Poisoning by unspecified narcotics, undetermined, sequela |
| T40.605 | Included miscode |
| T40.605A | Adverse effect of unspecified narcotics, initial encounter |
| T40.605D | Adverse effect of unspecified narcotics, subsequent encounter |
| T40.605S | Adverse effect of unspecified narcotics, sequela |
| T40.691 | Included miscode |
| T40.691A | Poisoning by other narcotics, accidental (unintentional), initial encounter |
| T40.691D | Poisoning by other narcotics, accidental (unintentional), subsequent encounter |
| T40.691S | Poisoning by other narcotics, accidental (unintentional), sequela |
| T40.692 | Included miscode |
| T40.692A | Poisoning by other narcotics, intentional self-harm, initial encounter |
| T40.692D | Poisoning by other narcotics, intentional self-harm, subsequent encounter |
| T40.692S | Poisoning by other narcotics, intentional self-harm, sequela |
| T40.693 | Included miscode |
| T40.693A | Poisoning by other narcotics, assault, initial encounter |
| T40.693D | Poisoning by other narcotics, assault, subsequent encounter |
| T40.693S | Poisoning by other narcotics, assault, sequela |
| T40.694 | Included miscode |
| T40.694A | Poisoning by other narcotics, undetermined, initial encounter |
| T40.694D | Poisoning by other narcotics, undetermined, subsequent encounter |
| T40.694S | Poisoning by other narcotics, undetermined, sequela |
| T40.695 | Included miscode |
| T40.695A | Adverse effect of other narcotics, initial encounter |
| T40.695D | Adverse effect of other narcotics, subsequent encounter |
| T40.695S | Adverse effect of other narcotics, sequela |

**MOUD Medication Formulations**

BUPRENORPHINE HCL-NALOXONE HCL 8-2 MG SL FILM

SUBOXONE 8-2 MG SL FILM

BUPRENORPHINE HCL-NALOXONE HCL 8-2 MG SL SUBL

BUPRENORPHINE HCL 8 MG SL SUBL

BUPRENORPHINE HCL-NALOXONE HCL 4-1 MG SL FILM

BUPRENORPHINE HCL-NALOXONE HCL 2-0.5 MG SL SUBL

BUPRENORPHINE HCL-NALOXONE HCL 2-0.5 MG SL FILM

SUBOXONE 2-0.5 MG SL FILM

BUPRENORPHINE HCL 2 MG SL SUBL

BUPRENORPHINE HCL-NALOXONE HCL 12-3 MG SL FILM

SUBOXONE 4-1 MG SL FILM

naltrexone (VIVITROL) 380 MG Recon Susp injection

naltrexone (VIVITROL) injection 380 mg

VIVITROL 380 MG Recon Susp injection

Naltrexone (VIVITROL IM)

naltrexone (FOR:VIVITROL) 380 MG Recon Susp injection

naltrexone (for:VIVITROL) injection 380 mg

NALTREXONE IM

NTX Injection (Print)

Sublocade 100 mg

Sublocade 300 mg

Probuphine Subdermal 74.2 mg
